# Fentanyl Purity and Overdose Decline: A Reexamination of Geographic Trends

**DOI:** 10.64898/2026.04.23.26351605

**Authors:** Nabarun Dasgupta, Adams Sibley, Paula Gildner, Katherine Gora Combs, Lori A. Post, Sam Tobias, Alex H. Kral, Rosalie Liccardo Pacula

## Abstract

Drug overdose deaths in the United States reached record levels during the fentanyl era before recently declining. A plausible hypothesis is that a sudden drop in fentanyl purity (“supply shock”) beginning in 2023 caused the downturn in overdose mortality. We evaluated this hypothesis by replicating a published analysis with regional overdose data, using models that account for time trends and autocorrelation, and negative control indicators to test for spurious correlation. Replicating the original paper, we extracted fentanyl purity data from a figure published by the Drug Enforcement Agency; overdose mortality was derived from standard vital statistics sources. We found that when fentanyl purity was rising, the national purity series did not track overdose increases in most regions and showed only a modest association in the West. When both purity and mortality later declined, the observed associations were also seen with unrelated macroeconomic indicators that shared the same time pattern. Canadian studies revealed variation in both strength and direction of association between fentanyl purity and overdose deaths at sub-province levels. Differential selection bias in law enforcement purity assessment may compromise longitudinal and geographic generalizability of DEA data, notably simulated reporting for higher purity wholesale samples in 2023-4. National fentanyl purity alone does not provide sufficient explanation for the recent decline in overdose deaths. A reduction in localized purity may influence overdose decreases, but other causes must also be considered, including drug supply changes, public health interventions, demographic shifts, behavior changes, inter-cartel dynamics, and fewer new initiates.

**Teaser:** Fentanyl purity alone does not explain the increase and decrease in overdose deaths across all regions of the United States.

## INTRODUCTION

Over the last half-decade, the United States experienced record drug overdose mortality, driven primarily by unregulated fentanyl, followed by an unprecedented decline (*1–5*). As of July 2025, all states showed substantial declines from peak overdose, of at least 19%, with some states approaching 50% decline from peak (*6*). Vangelov et al. (*7*) proposed that a fentanyl “supply shock” beginning in May 2023, precipitated by Chinese government actions, was the primary driver of this decline.

Statistically, the conjecture hinges on their observation that starting March-to-July 2023, powder purity appeared to fall from around 25% to 11% by the end of 2024. And that fentanyl overdose deaths declined by “more than half” from a national peak in May 2023 during the same time. They offer a correlation statistic (r = 0.37) for the strength of the association between January 2019 to October 2024 (*7*).

Others have proposed a similar hypothesis in Canada (*8*). Given regional variation in overdose rates (*9*, *10*) and subgroup trends (*11*, *12*), we tested whether the “supply shock” hypothesis holds at sub-national levels in North America. We were motivated by the fact that overdose death rates by state peaked as early as 2021, with some declining gradually (*6*). Broadly summarizing previous drug cycles, rapid drops in overdose are believed to more likely represent drug supply disruption, such as that which occurred following the heroin drought in Australia (*13*). Gradual declines suggest structural factors (*14*) including interventions, behavior change, and long-run demographic shifts. This distinction has important policy priority-setting implications, informing resource allocation and intervention design. If a purity-only hypothesis can explain the recent decline in overdose, the role of interventions, behavior change, and demographics becomes less salient. Conversely, if purity can only explain a limited portion of overdose declines, the importance of interventions is heightened. Therefore, testing the robustness of this hypothesis has considerable implications for public health, implementation science, and government policy.

### Population Studies of Overdose Increase

Our main test was whether overdoses increased in tandem with fentanyl purity before decline. We analyzed two periods: Block 1 (July 2022–September 2023), when purity rose, and Block 2 (October 2023–September 2024), when purity declined. Prior studies reported positive associations between fentanyl supply measures and overdose mortality at state/province and national levels (*15–18*), though some appeared inflated by autocorrelation (*17*).

Quantifying the exact nature of the association is benefitted by understanding geographic variation in drug purity (*19*), variations in where and when fentanyl initially emerged (*20–22*), adulterants (*22–25*), and route of administration (*26*). A previous study in British Columbia found routine two-fold fluctuation in fentanyl purity over three years (*27*). Even within cities, retail fentanyl purity varies widely; in Los Angeles (*28*), expected “fentanyl” samples ranged from <1 mg to 697 mg active ingredient per gram, with median purity 10.0%.

To test the robustness of the supply shock hypothesis, we replicated the Vangelov et al. study at a sub-national level. We also used negative controls using macroeconomic indicators to evaluate the presence of shared time trend structures.

### Methods Limitations of Original Analysis

#### Trend and Autocorrelation

Purity and mortality series showed autocorrelation and nonlinearity, so omnibus correlation statistics would be inflated and violate independence assumptions. The original analysis did not specify which correlation coefficient was used to describe the strength of association, the associated assumptions, nor how statistical significance was assessed, limiting replicability. We therefore used ARIMA-based transfer function models (*29*) and two negative controls to isolate associations beyond shared secular time trends.

#### Aggregation Bias

The published analysis (*7*) was conducted at the aggregated national level, despite local variability in both fentanyl purity and overdose (*30*). The explicit assumption was that both death rates and fentanyl purity were nationally distributed uniformly. The unregulated drug supply can be highly regionalized, and many states had declining overdose rates (*20*) years before the onset of the purported supply shock. We were concerned that the independent (purity) and dependent (fatal overdose rate) variables could be subject to aggregation bias in the form of Simpson’s Paradox (*31*), wherein the aggregate national rate increases (despite declines in some states) because the national rate is a weighted composite that can be driven upward by larger or faster-growing increases in a subset of states, reversing or obscuring regional trends. Therefore, using a common-input, differential-response design, we compared overdose mortality rates by four US Census Regions to the national fentanyl purity time series, focusing on two temporally adjacent periods of increasing and decreasing fentanyl purity comprising the crux of the supply shock hypothesis.

## RESULTS

### Descriptive Findings

Figure 1 shows that national fentanyl overdose mortality rose sharply beginning in Spring 2020, remained elevated through mid-2021, and began declining in mid-2023. DEA-reported powder purity (Supplement S1) followed a different trajectory, rising mainly from 2022 through 2023 before declining in 2024.

**Fig. 1.**
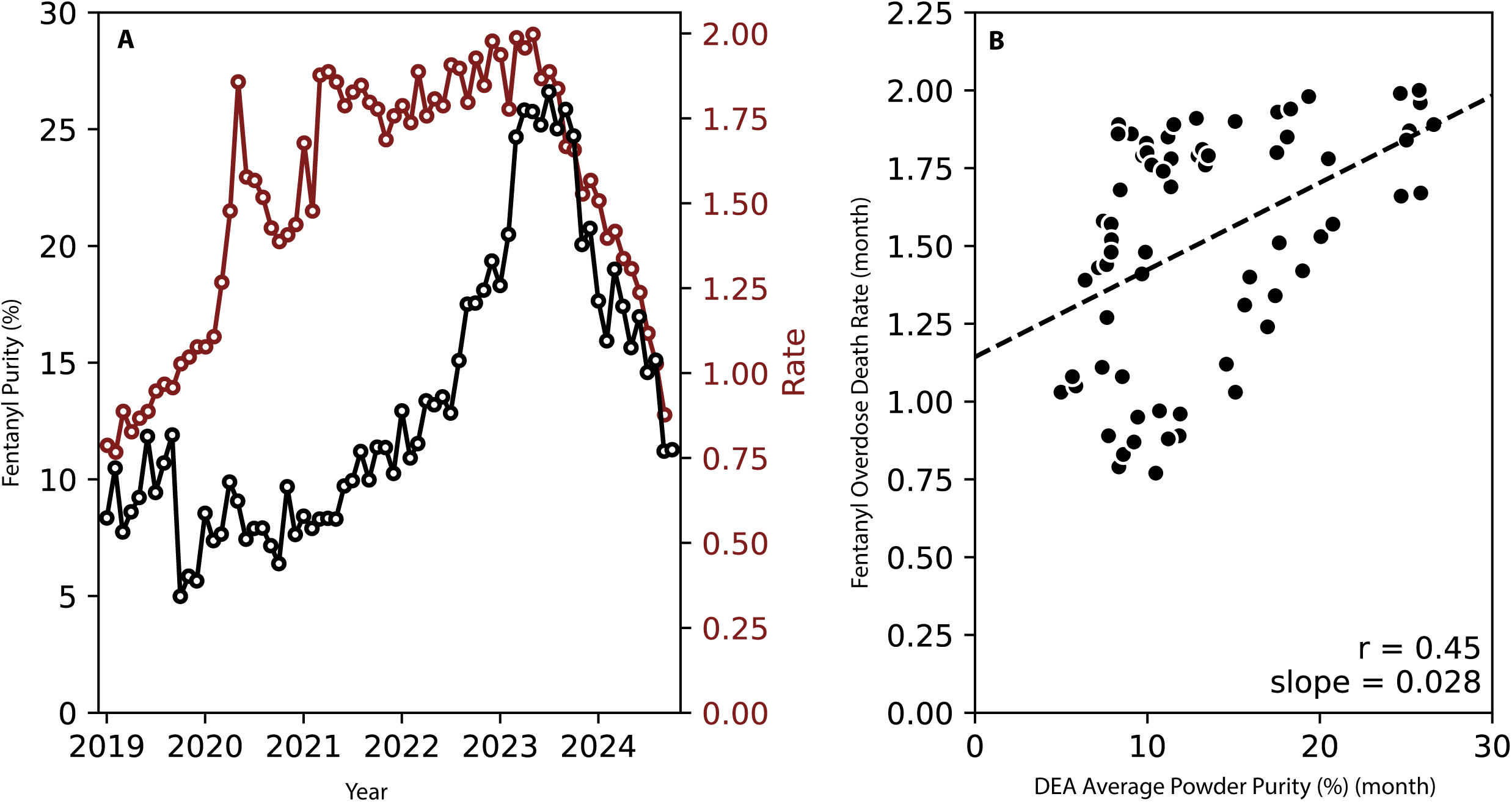
Fentanyl Overdose Mortality and Fentanyl Purity. Panel A: Time series of DEA fentanyl powder purity (%) plotted against the US overdose death rate (per 100,000) on a shared time axis (January 2019 to October 2024) with separate y-axes. Powder purity (left y-axis) is shown in black; the monthly fentanyl overdose death rate (right y-axis) is dark red. Panel B: Scatterplot of monthly overdose death rate versus powder purity, with a fitted least-squares regression line (dashed).

**Fig. 2.**
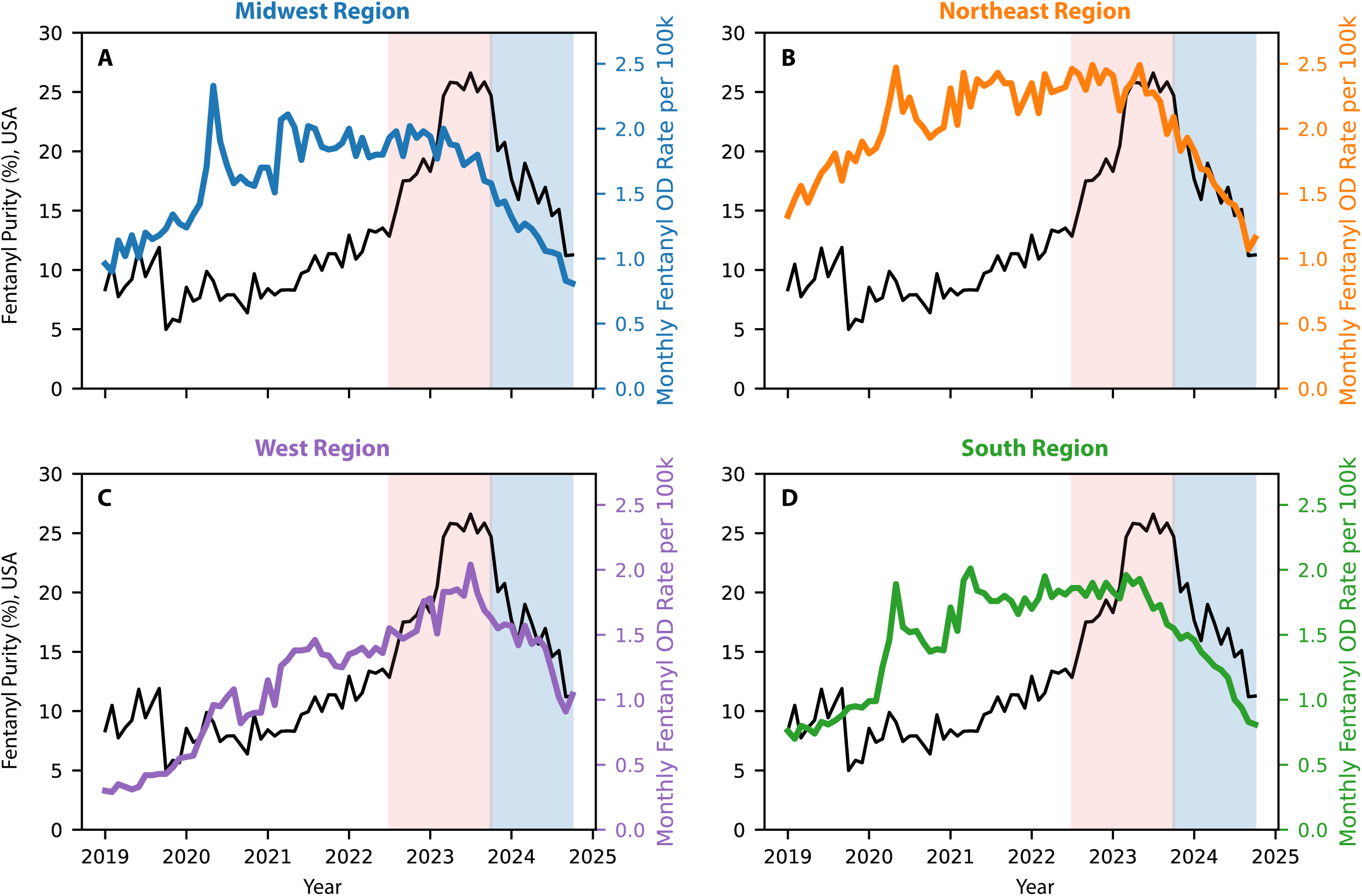
Fentanyl Overdose Mortality and Fentanyl Purity by US Census Region. Monthly fentanyl powder purity (black line, left axis; national DEA estimate) and fentanyl overdose mortality rate per 100,000 (colored lines, right axis) by US Census region, January 2019–October 2024. Shaded bands indicate the two analytic blocks: Block 1 (pink; July 2022–September 2023, rising purity) and Block 2 (blue; October 2023–September 2024, declining purity). The purity series is identical across all four panels, reflecting its status as a single national estimate without geographic stratification. Note divergent trajectories of purity and mortality in the Northeast (**B**), Midwest (**A**), and South (**D**) during Block 1, contrasting with the West (**A**) where both series co-move.

Raw Pearson correlations between national purity and mortality were negative for the Northeast (raw r = −0.47), Midwest (raw r = −0.54), and South (raw r = −0.27), leading to the improbable appearance that overdoses *declined* while fentanyl purity *increased*. Hence, accounting for autocorrelation and geographic stratification are essential to interpreting the impact of the purported supply shock.

At the start of Block 1 (July 2022) “national” fentanyl powder purity was 12.8% (Table 1), doubling to 25.8% by September 2023. Fentanyl overdose mortality rates decreased substantially during that same time period, with the greatest drop rate in the Midwest (Δ-45.4%), only increasing slightly (9%) in the West.

**Table 1.** Fentanyl powder purity and overdose rates during two blocks of purity change.

| Block 1 (July 2022 to September 2023) |  |  |  |  |  |  |
| --- | --- | --- | --- | --- | --- | --- |
|  | DEA* Fentanyl Powder Purity |  |  | Monthly Fentanyl Overdose Death Rate (per 100,000 population) |  |  |
| Region | July 2022 | September 2023 | Percent Change | July 2022 | September 2023 | Percent Change |
| Northeast | 12.8% | 25.8% | +101.5% | 2.46 | 1.96 | -20.3% |
| Midwest | 12.8% | 25.8% | +101.5% | 2.93 | 1.60 | -45.4% |
| South | 12.8% | 25.8% | +101.5% | 1.86 | 1.58 | -15.1% |
| West | 12.8% | 25.8% | +101.5% | 1.55 | 1.69 | +9.0% |
| Block 2 (October 2023 to September 2024) |  |  |  |  |  |  |
|  | DEA Fentanyl Powder Purity |  |  | Monthly Fentanyl Overdose Death Rate (per 100,000 population) |  |  |
| Region | October 2023 | September 2024 | Percent Change | October 2023 | September 2024 | Percent Change |
| Northeast | 24.7% | 11.2% | -54.6% | 2.09 | 1.07 | -48.8% |
| Midwest | 24.7% | 11.2% | -54.6% | 1.58 | 0.83 | -47.5% |
| South | 24.7% | 11.2% | -54.6% | 1.55 | 0.83 | -46.5% |
| West | 24.7% | 11.2% | -54.6% | 1.63 | 0.91 | -44.2% |

\* DEA, Drug Enforcement Agency

During Block 2 fentanyl purity declined (Δ-54.6%) from 24.7% in October 2023 to 11.2% in September 2024. Monthly fentanyl overdose death rates declined, 44.2% to 48.8% across Census Regions during Block 2.

### National Transfer Function Models

We first replicated Vangelov et al.’s analysis using autoregressive-corrected models for the sake of direct comparison to the publication. (This analysis was not included in the pre-registration but requested during the manuscript review process.) TFM was applied to national overdose mortality against the national purity series, the full-period gain was statistically significant but weak (gain = +0.026 deaths per 100,000 per percentage-point change in purity, p = 0.011, TFM R² = 0.249); however, residual autocorrelation was detected in the full-period model (Ljung–Box p = 0.001), consistent with a structural break between the rising and declining phases and supporting the analytic decision to estimate the two blocks of increase/decline separately.

Dividing the full period into its two structurally distinct blocks reveals that this pooled national association is driven primarily by Block 2, where the gain approached but did not reach conventional significance (gain = +0.022, p = 0.059, TFM R^2^=0.38). The Block 1 gain (the critical test of whether *rising* purity tracked rising mortality) was not meaningful (gain = +0.034, p = 0.143, R² = 0.184). This block-stratified result points to the purity–mortality association detected in national data reflecting convergent post-2023 decline trajectories rather than consistent pharmacological coupling across both increase and decline phases. While these analyses afford a general sense of decoupling of the purity-overdose relationship nationally, they are prelude to the *a priori* analyses where regional models were central to testing the purity shock hypothesis.

### Regional Models: Period of Fentanyl Purity Increase

In Block 1, during the period of purported rising fentanyl purity (July 2022 to September 2023), the transfer function gain (Table 2) was substantively meaningful only in the West (gain = 0.056 deaths per 100,000 per month per percentage-point change in powder fentanyl purity, p = 0.10, TFM R² = 0.22). The Northeast and Midwest showed near-zero gains (0.008 for both, p > 0.80, TFM R² = 0.005), indicating that month-to-month innovations in the “national” purity series carried no predictive information for mortality rates. The South showed similarly weak association (gain = 0.012, p = 0.59). The discordance between TFM R^2^ and penalized R^2^ further confirms that the fentanyl purity series in the Northeast, Midwest, and South make no meaningful information gains on explaining overdose mortality rates.

**Table 2.** Transfer function model results for the association between fentanyl purity and monthly fentanyl overdose mortality rate by US Census region.

| Region | n | Gain | SE | p | TFM R<sup>2</sup> | Penalized R<sup>2</sup> | Raw r |
| --- | --- | --- | --- | --- | --- | --- | --- |
| Block 1 (July 2022 to September 2023) |  |  |  |  |  |  |  |
| Northeast | 15 | +0.008 | 0.031 | 0.815 | 0.005 | -0.085 | −0.47 |
| Midwest | 15 | +0.008 | 0.032 | 0.810 | 0.005 | -0.085 | −0.54 |
| South | 15 | +0.012 | 0.021 | 0.592 | 0.027 | -0.061 | −0.27 |
| West | 15 | +0.056 | 0.032 | 0.103 | 0.223 | 0.153 | +0.79 |
| Block 2 (October 2023 to September 2024) |  |  |  |  |  |  |  |
| Northeast | 12 | +0.027 | 0.011 | **0.038\*** | 0.436 | 0.366 | +0.92 |
| Midwest | 12 | +0.021 | 0.007 | **0.019\*** | 0.519 | 0.459 | +0.95 |
| South | 12 | +0.010 | 0.006 | 0.161 | 0.230 | 0.134 | +0.86 |
| West | 12 | +0.020 | 0.015 | 0.226 | 0.177 | 0.074 | +0.83 |

**Abbreviations:** SE, standard error. Gain represents the transfer function steady-state multiplier: the estimated change in monthly mortality rate per 100,000 associated with a sustained 1 percentage-point change in national fentanyl purity, after prewhitening both series via an ARIMA(1,1,0) filter applied to the input (purity). TFM = Transfer function model. Raw r = Pearson correlation between purity and mortality in levels (unprewhitened). R<sup>2</sup> = variance explained by the transfer function model on prewhitened series. Penalized R<sup>2</sup> = R<sup>2</sup> adjusted for number of predictors and sample size, providing a more conservative measure of fit that decreases when predictors do not improve explanatory power. \* p < 0.05.

### Regional Models: Period of Fentanyl Purity Decrease

The purity-overdose association for the West (TFM R^2^ = 0.18, p=0.226) was not meaningful in Block 2 (October 2023 to September 2024), despite being plausibly correlated in Block 1. This divergent finding casts doubt on the ability of the purity series to explain overdose rates in the West. Stated another way, it is difficult to conclude that overdoses were declining *only* due to fentanyl purity in Block 2 because overdoses rates were decoupled from increasing purity in Block 1. For the South, the purity-mortality relationship remained weak in both time periods. This region includes southern Appalachian states with entrenched, elevated intergenerational overdose rates.

In Block 2, as both purity and mortality declined, transfer function gains became statistically significant in the Northeast (0.027, p = 0.04) and Midwest (0.021, p = 0.02), with adjusted R² values explaining variance of 0.44 and 0.52, respectively. This apparent strengthening of the purity-mortality relationship may reflect convergent macro time trends, where both series decline simultaneously along with broader post-COVID societal indicators, rather than genuine validation of the purity series as a causal predictor of mortality, necessitating the use of negative controls.

### Negative Controls

To assess spurious correlation, we compared overdose mortality with two macroeconomic indicators (retail price inflation and consumer sentiment, Supplement S2) that have no direct pharmacologic pathway to overdose death.

In unadjusted time-series analysis (Table S3), overdose mortality was strongly correlated with both inflation (Pearson raw r = +0.63, R² = 0.40) and consumer sentiment (raw r = −0.73, R² = 0.53), associations of comparable or greater magnitude to the purity-mortality correlation reported in the original publication. Raw cross-correlation between overdose mortality rate and each economic indicator (Fig. S3, panel C) would have shown “statistically significant” associations at any month between lags of −12 to +12 months, further evidence that the purity-overdose association is spurious.

Following adjustment for autocorrelation and time trend, both associations reversed direction entirely: the overdose-inflation correlation shifted from r_raw_ = +0.63 to r_TFM_ = −0.34, and overdose-sentiment correlation from r_raw_ = −0.73 to r_TFM_ = +0.32. Sign reversals of this magnitude are diagnostic in TFM models; when prewhitening transforms a strong positive correlation into a negative one, the original association is, by definition (*29*), an artifact of shared temporal structure rather than any causal underlying relationship. Fentanyl purity follows the same post-COVID arc as these macroeconomic indicators, rendering purity structurally indistinguishable from unrelated series. The negative control experiments reveal that the hypothesized purity-overdose associations cannot be interpreted as causal and are more likely spurious.

### Comparison with Canada

As Vangelov et al. did, we also compared our findings to British Columbia, where Tobias et al. (*15*) used geographically resolved fentanyl concentration data from community drug checking services to model the same fentanyl purity-mortality relationship, using generalized additive mixed models stratified by n=16, sub-province health districts). Each one percentage-point increase in fentanyl concentration was associated with a +0.072 gain in drug-related mortality per 100,000 population (p = 0.029), an effect size roughly two to three times larger than our observed strongest transfer function gains in Block 1.

Critically, the British Columbia study also found significant regional heterogeneity in the strength of association, with gain estimates ranging from −0.024 to +0.171 across health service delivery areas, indicating that even with geographically specific data, the purity-mortality relationship varies highly by local context. The attenuation of our effect sizes relative to those reported by Tobias et al. is consistent with measurement error introduced by geographic mismatch between the exposure and outcome.

In Toronto, opioid mortality peaked in late 2020 through early 2021 and subsequently entered a sustained downward trajectory, albeit with a prolonged period of fluctuation before sharper declines observed in 2024. In contrast, mean fentanyl purity remained elevated through 2023 and early 2024 before declining (*32*). This lack of temporal alignment suggests that reductions in average potency may have contributed to later stages of the trajectory but are unlikely to explain the initial downward inflection in mortality.

## DISCUSSION

Our primary test for assessing the supply shock hypothesis was to evaluate whether increases in the fentanyl purity were associated with increases in overdose mortality. Without proper time-series adjustment for trend and autocorrelation, correlations during the period of purity increase would have found a *negative* association in most of the country (raw Pearson r = −0.54 in the Midwest, raw r = −0.47 in the Northeast), suggesting that *higher purity coincided with fewer deaths* paradoxically. Previous analyses (*15*, *18*, *30*, *33*, *34*) have shown a strong positive correlation between fentanyl purity and overdose mortality; if we could not replicate even this established association in Block 1, it seems the purity series published in the National Drug Threat Assessment should be used cautiously by scientists in making causal inference. DEA cautioned (*35*) against exactly this interpretation due to temporary sampling directives in 2023-4 that focused on higher purity wholesale samples (800 grams or more).

In time series analysis, when multiple social indicators move in the same direction, associations can arise spuriously. Our negative control analyses show that Block 2 overdose rate associations are compatible with shared post-COVID time trend structure rather than a specific pharmacologic effect of fentanyl purity. Contemporaneous declines in overdose mortality from psychostimulants and non-opioid substances (*36*) lend further credence to considering broader societal time trends as influencing overdose mortality declines.

After correcting for assumption violations, the national purity series was meaningfully associated with overdose increases only in the West during Block 1, and even there modestly. In the Northeast and Midwest, the same series explained essentially none of the month-to-month variation during the rise phase, then became only mildly associated during the decline phase. Together, these findings support two conclusions: the purity series is not nationally representative, and structural factors likely contributed materially to both the rise and fall in overdose mortality. The South further illustrates this mismatch: despite heavy representation in DEA field-lab sampling (44% of all samples in 2023, Supplement S3), the purity series showed weak association with overdose in both blocks. This likely reflects the fact that several southern states had already entered decline (*6*) years before the purported 2023 supply shock. Some local associations may still exist, but they would require geographically resolved data to interpret credibly. A single national purity series regressed against national mortality is insufficient to support causal claims about a fixed-time-point supply shock in a large, heterogeneous country.

These findings have practical policy implications. If no clear national supply shock explains overdose downturns, greater attention should be paid to local structural factors that changed over the same period, including naloxone diffusion, treatment expansion, recovery supports, and demographic shifts (*1*). We do not foreclose the possibility that fentanyl purity declined in specific locations around 2023, including settings where fentanyl was displaced by sedatives or numbing agents (*37*, *38*). However, a simplistic supply shock is not supported by the evidence.

We also uncovered that the purity series used in the DEA figure does not align with other DEA reporting. For example, in 2024, the DEA reported (*39*) national powder fentanyl purity of 11.3%, whereas the extracted data from the figure had it as 15.2%. In 2023, the reported average DEA field lab powder purity was 19.7% and DEA’s Fentanyl Profiling Program purity was 33.5% (*35*), whereas figure-extracted data had it as 23.3%. These values were substantially higher than contemporaneous retail-level estimates from Los Angeles (*28*), raising concern about representativeness. We emphasize the need for caution when using poorly documented enforcement-derived data in policy analyses, and sampling methodology and datasets should be explicitly disclosed as part of governmental reports.

### Localized Purity Impacts

Despite our analysis not being able to support a supply shock hypothesis at national or regional levels, we contend that local variation in fentanyl supply is still likely to be real and consequential. An arising question is what the appropriate level of geographic granularity would be to interrogate the purity-overdose association with fidelity. This question is consistent with our original suspicion of Simpson’s Paradox, whereby aggregate fentanyl purity and overdose rates mask local variation.

Returning to Tobias et al. (*15*), the most geographically resolved evidence from British Columbia reinforces the need to precisely measure local conditions of the exposure, namely fentanyl concentration at the retail level. Point-of-care drug-checking samples are arguably the closest available approximation to true street-level exposure, in contrast to wholesale law enforcement seizures. Median fentanyl concentration varied across districts in 2023, from 6.0% on Central Vancouver Island to 18.2% in North Shore/Coast Garibaldi, a more than three-fold difference. Tobias et al. also note that areas with more urban communities carried higher fentanyl concentrations than rural ones, but the distribution was not uniform.

With such wide heterogeneity in the exposure, it is not surprising that the concentration–mortality association in Tobias et al. was likewise heterogeneous. A random-slope specification was statistically required to model the association, isolating and allowing each sub-province health district to have its own trajectory. Even despite this, the resulting model’s explanatory power ranged from R² = 0% to 58% across areas. Because the strongest slopes appeared in both a dense urban setting (Vancouver, β = 0.169) and a largely rural one (Northern Interior, β = 0.171), the relationship is shaped by local context in ways that do not reduce to a simple urban–rural dichotomy. This contingency is critical, pointing beyond simple pharmacological explanation, and requiring consideration of local structural factors such as market maturity, time since fentanyl emergence, and hyperlocal drug distribution networks. The assumption of uniformity in the single national supply shock hypothesis ignores this evidentiary nuance.

In the British Columbia study, estimate precision was greatest in populous districts with dense drug checking coverage and least stable where sampling was sparse, so the apparent strength of a purity–mortality association depends on observing the local drug supply accurately. The national series we analyzed is built on sampling that is profoundly uneven across US states and regions. As detailed in Supplement S3, 44% of 2023 DEA powder exhibits originated in the South (n = 3,370) but only 14% in the West (n = 1,117), of which California alone accounted for 59% (n = 657). By contrast, states like Alaska contributed only n=18. Placing the Vangelov et al. analysis in the context of our analysis, the regional signal was decoupled from where it appears to perform: the West, where the series tracked mortality most closely, rests on the sparsest and most geographically concentrated sampling base. In contrast, the South, which dominates the national sample, showed weak association in both blocks. A purity figure aggregated from such unequal representation cannot be read as equally informative everywhere, and a weak association at regional or aggregate scales may reflect the sampling structure of the exposure measure as much as any true local effect. Resolving this question will require localized, adequately powered exposure data, such as those generated by drug checking interventions (*40*).

### Alternative Causal Hypotheses

The basis for the supply shock hypothesis is the pharmacological expectation that higher fentanyl concentrations should increase risk of respiratory depression and fatal overdose at the individual level. However, comparing opioid-naïve individuals to those with opioid tolerance from chronic use, experimental studies have shown fentanyl to require a 4.3-fold difference in concentration to achieve the same 50% ventilatory depression in both populations (*41*). Therefore, population-level heterogeneity in the relationship between purity and overdose may be conditional on local levels of opioid tolerance, similar to the protective effect of steady state pharmacokinetics during methadone maintenance (*42*). We foregrounded the opioid-tolerance hypothesis first not because it outranks other explanations, but because it is the mechanism most directly continuous with the pharmacological premise of the supply shock hypothesis itself. Beyond the purity consideration, we posit five non-exclusive mechanisms that may explain the decline in overdose.

#### Drug Supply Changes

Two sequential waves of illicit drug supply changes are suspected to have broadly modify overdose risk. First, the replacement of heroin with fentanyl, spreading east to west, catalyzed an unprecedented acceleration in overdose deaths (*20*, *36*), with state-by-state (*43*) trajectories trending towards baseline in a consistent multi-year cascade. Second, in places where fentanyl had appeared earliest, the unregulated drug supply then become adulterated with strong sedatives and numbing agents, accompanied by reports of growing user dissatisfaction (*44*), conditions that can suppress consumption, both independent of and as a reaction to fentanyl concentration (*45*, *46*). Adulteration of fentanyl with xylazine has been shown to result in compensatory protective behavior to use less drugs or less frequently (*44*), and less severe overdose in animal models (*47*) and medical practice (*48*). The upstream causes of the drug supply fluctuations are poorly understood, but dynamics within and between organized criminal organizations (*49*) cannot be discounted, with external pressure exerted from interdiction.

#### Demographic Shifts

Second, demographic shifts were evident during the study period, which cannot be explained by a simple national supply shock hypothesis. The population of people who use opioids has been shifting in recent years, with an established cohort ageing through middle age and fewer new initiates entering use (*36*, *50*), structurally reducing the size of the at-risk population over time. During the study period, as overdose deaths reached peak levels in the United States, the greatest growth (*51*, *52*) in fentanyl mortality was observed in African-American and Native American men. A supply shock hypothesis cannot cohesively explain these shifting disparities.

#### Behavior Changes

Within drug using networks, behavioral adaptation, using smaller amounts, using less often, using test strips, shifting away from injection towards smoking and snorting (*53*, *54*), and adopting safer drug use practices have been shown to alter overdose risk (*55*, *56*). The aforementioned length of time elapsed since the emergence of unregulated fentanyl in the local drug supply may afford people time to adapt to the different potency, while at the same time causing an initial rapid rise in overdose deaths (*43*) that diminish the pool of current illicit drug users, in turn creating intergenerational negative feedback loops (*57*, *58*) that discourage continued illicit drug use and reduce new initiates.

#### Public Health Interventions

Community-based and clinical interventions expanded substantially over the study period, particularly since the height of the COVID pandemic, including naloxone distribution (*43*) and broadened access to evidence-based treatment (*1*, *59*). In addition, distribution of opioid settlement funds, reduced stigma for seeking treatment among young adults and youth (*60*), and policy changes (*61*) including telehealth and Medicaid expansion (*62*), further created the conditions for fundamental changes in overdose risk by creating off-ramps for problematic substance use. While these interventions may take time to make meaningful impact on overdose rates, the simultaneous implementation of these many courses of action could deliver faster results than each would be expected to produce in isolation.

#### Societal Post-Pandemic Recovery

There is mounting evidence (*63*) that overdose declines parallel post-COVID pandemic recovery trends seen in broader societal indicators, including violent crime (*64*, *65*), motor vehicle fatalities (*66*), and suicide (*67*). These downward trajectories are similar to the macroeconomic series examined in our negative control analysis. Our finding that the purity–mortality association rides on shared post-COVID temporal structure with unrelated phenomena suggests that broad societal trends cannot be entirely disentangled with the decline in overdose deaths, although their causal connection remains challenging to establish.

#### Limitations

Neither our study, nor others (*7*, *18*, *19*, *30*, *68*), accounted for myriad interventions implemented to reduce overdose mortality at a local level. These overlapping efforts are notoriously difficult to measure prospectively over large areas, as we previously (*69*) demonstrated.

We acknowledge that the analytic blocks were short for transfer function modeling. That constraint was imposed by the brief rise-then-fall structure of the purity series; pooling across the structural break would have violated stationarity assumptions. We therefore used a parsimonious ARIMA(1,1,0) specification and BIC-based model selection. The consistency of null findings across regions in Block 1 supports the robustness of the central result despite limited power. Time-varying under-ascertainment of fentanyl’s involvement in overdose, especially in jurisdictions where postmortem toxicology practices changed over time, may have affected regional mortality estimates (*70*).

The purity series was manually digitized from a figure and may contain measurement error. We did not model adulterants such as xylazine, medetomidine, or bromazolam, which may alter both overdose risk and compensatory behavior; fentanyl purity alone is an incomplete measure of mortality risk in the contemporary synthetic drug supply.

We acknowledge that systematically collected sub-national purity data would have been a boon to this analysis, and the common input-differential response design is not inherently causal. Without more granular governmental or drug checking data, a more directly causal modeling was not possible, and our intent was to evaluate the hypothesis posed by Vangelov et al. through replication.

For data derived from drug checking services in British Columbia, we also note that different health services areas had varying levels of sampling volume (*15*). We are not aware of any precedent to accurately draw a fully representative sample of the unregulated drug supply over large geographic areas (*40*).

We also did not examine demographic shifts in overdose mortality, including age and race/ethnicity, which may be central to explaining the recent decline. A fundamental causal explanation for the decline in overdose deaths is likely fewer new initiates to illicit opioid use (*14*). These limitations should be addressed in future studies.

#### Methodological and Ethical Implications

Methodologically, national time series should not be entered into regression models without addressing autocorrelation, time trends, and sub-national variation. And enforcement-derived data should not be used without adequate documentation of sampling.

We do not dispute that drug supply characteristics and overdose mortality are linked. Our point is narrower: this national purity series does not provide causal proof. In the current synthetic-drug era, “concentration” may be a more useful concept than “purity,” because mortality risk depends on a changing mixture of fentanyl, sedatives, novel opioids, and other co-occurring substances. A decline in fentanyl purity alone may not correspond to a decline in total overdose risk.

No statistical model can substitute for the obligation to provide humane, effective care to reduce overdose deaths. The geopolitical framing of the supply shock hypothesis risks implying that interdiction was the sole causative factor, but the recent history of drug supply changes, interventions, demographic shifts, and behavior changes require us to consider more complex and interwoven explanations. A purely reductionist approach demeans the considerable efforts enacted across the United States and Canada to reduce overdose deaths, and we entreat researchers to engage in pursuing analysis of multi-cause possibilities.

## MATERIALS AND METHODS

### Data Sources

#### Fentanyl purity

DEA publicly reports purity quantification from regional field labs where law enforcement seizures (ostensibly at or near street retail level) are quantified for prosecutorial evidence. In addition, a smaller number of intentionally selected samples are sent to the DEA’s Fentanyl Profiling Program (FPP) In 2023, the focus of both shifted to higher volume (“wholesale”) drug seizures of greater than 800 grams (*35*), with average powder purity via DEA field labs as 19.7% and selected higher volume samples via FPP as 33.5%. Therefore, the data used by Vangelov et al. from the National Drug Threat Assessment may represent a sampling bubble in 2023-4, where higher purity samples were comingled with retail samples. We recreated the purity series used by Vangelov et al., extracting data points from the fentanyl powder figure in the 2025 DEA National Drug Threat Assessment (*71*) using code-assisted manual coordinate extraction to handle pixel-level anti-aliasing, Supplement S1. We were able to replicate (Fig. S1) the fentanyl powder purity time series with fidelity: Lin’s concordance correlation coefficient for reproducibility was 0.999.

#### Overdose Data

Monthly drug overdose mortality counts were obtained from OD Pulse, an overdose surveillance data platform maintained by Northwestern University (*36*). OD Pulse pre-processes mortality data from the CDC Wide-ranging Online Data for Epidemiologic Research (WONDER). Overdose deaths were defined X40-X44, X60-X64, X85, or Y10-Y14 (*36*, *72*), and coded as T40.4 (synthetic opioids, predominantly unregulated fentanyl and analogues). Monthly death counts were aggregated by the four US Census regions (Northeast, Midwest, South, West) and converted to population-adjusted mortality rates per 100,000 using Census Bureau intercensal population estimates.

### Statistical Models

We specified a common-input, differential-response design: the same national purity series served as the exposure for all four Census Regions, with region-specific monthly overdose mortality rates per 100,000 as the output. This design directly tests the assumption underlying Vangelov et al.’s analysis, that a single national purity measure is equally informative across the country, by using the original study’s own premise as the testable hypothesis. If the national series were truly representative of local supply conditions, we would expect comparable transfer function gains across regions. Conversely, divergent results constitute evidence of geographic mismatch between the national measure and local supply.

To assess the temporal relationship between fentanyl powder purity and overdose mortality rates across Census Regions, we estimated TFMs following the Box-Jenkins procedure (*29*, *73*). We first fit an ARIMA time-series model that captures both trend and month-to-month momentum in the purity series, with residuals representing each month’s value that could not have been predicted from the series’ own history. Applying the identical filter to the mortality series removed shared temporal structure, with surviving correlation reflecting the genuine contemporaneous relationship between changes in purity and changes in mortality. This conservative procedure preserves the true purity-mortality relationship and eliminates shared time trend (*29*).

The study period was divided into two analytic blocks: Block 1 (July 2022 to September 2023, n=15 months), characterized by rising fentanyl purity, and Block 2 (October 2023 to September 2024, n=12 months), characterized by declining purity. Model fit was assessed using adjusted coefficient of determination (R²), penalized for model complexity (*74*).

### Transfer Function Models

To assess the temporal relationship between fentanyl powder purity and overdose mortality rates across US Census Regions, we estimated transfer function models (TFMs) following the procedure established by Box-Jenkins (*29*, *73*), and applied in infectious disease surveillance (*75*) and other health fields (*76*). TFMs were applied to the primary analysis and for the negative controls. We first fit a time-series model that captures the trend and month-to-month momentum in the first series (purity), then extract the residuals, which are the component of each month’s value that could not have been predicted from the series’ own history. Applying the identical filter to the mortality series removes the same temporal structure from both, leaving only the direct monthly correlation (“innovations”) between purity and overdose, independent of prior trajectory. Any correlation that survives this filtering reflects a genuine contemporaneous relationship between changes in purity and changes in mortality, rather than an artifact of shared trends. Critically, this is a conservative procedure: if a true purity-mortality relationship exists in these data, prewhitening preserves it; if the correlation is driven entirely by shared trends, prewhitening eliminates it (*29*).

The input series was monthly fentanyl powder purity (percent) derived from the extracted DEA figure, which lacked geographic stratification. We specified a common-input, differential-response design: the same national purity series served as the exposure for all four Census Regions, with region-specific monthly overdose mortality rates per 100,000 as the output. This design directly tests the assumption underlying Vangelov et al.’s analysis, that a single national purity measure is equally informative across the country, by using the original study’s own premise as the testable hypothesis. If the national series were truly representative of local supply conditions, we would expect comparable transfer function gains across regions. Conversely, divergent gains constitute evidence of geographic mismatch between the national measure and local supply dynamics.

Based on visual time series inspection, the study period (July 2022 to September 2024; 27 months) was divided into two analytic blocks to reflect an apparent structural purity shift: Block 1 (July 2022 to September 2023, n=15 months), characterized by rising purity, and Block 2 (October 2023 to September 2024, n=12 months), characterized by declining purity. Models were estimated separately for each region-block combination (8 cells).

#### ARIMA-based Prewhitening

For each cell, we first prewhitened the input series by fitting an autoregressive integrated moving average (ARIMA) model to the purity series and extracting the residual innovations (*29*). Innovations are the residual component of each observation after removing predictable structure (trend and autoregressive dependence), representing the new information contributed by each time point that is orthogonal to the series’ own history. We selected ARIMA(1,1,0) as the prewhitening filter because the purity series exhibits both a clear trend (addressed by the differencing term, d=1) and modest persistence in month-to-month changes (addressed by a single autoregressive term, p=1), while the short series length (12–15 months per cell) precludes more complex specifications without overfitting. Bayesian Information Criterion (*77*) (BIC) comparison of lag-0 month versus lag-0,1 transfer function specifications confirmed that no additional complexity was warranted in any of the eight region-block cells. The same autoregressive-differencing filter was then applied to the mortality series, yielding a pair of prewhitened series free of shared autocorrelation structure. The same process was applied to the negative controls.

#### ARIMA Diagnostics

For each Region-Block pair (n=8), ARIMA fit was assessed for normality (e.g., Jarque–Bera) and serial correlation (e.g., Ljung–Box) (*78*). Visual inspection included residuals over time, residual autocorrelation function (ACF), and residual partial (PACF) (with 6-month lags), quantile-quantile (QQ)-plot of residuals, and plot of observed versus expected purity values over time.

#### Regression Model

We estimated impulse-response weights by regressing the filtered mortality series on the contemporaneous filtered purity series (or negative controls) via ordinary least squares. Impulse-response weights estimate magnitude of the mortality response to a one-unit “impulse” (innovation) in fentanyl purity at each month lag. In our lag-0 specification, a single weight represents how much of an “innovation” in purity passes through to mortality within the same month.

The primary interpretive metric is transfer function gain, defined as the sum of impulse-response weights, representing the expected change in monthly mortality rate per 100,000 population associated with a one percentage-point change in fentanyl purity. We report gain estimates with standard errors and significance levels for contemporaneous effect.

#### Assessing Model Fit via Penalized R^2^

We assessed model fit in the prewhitened domain using the coefficient of determination (R²). R² was computed as 1 − (SSR/SST), where SSR is the residual sum of squares from the prewhitened regression and SST is the total sum of squares around the mean of the prewhitened outcome. Because R² increases monotonically with additional predictors, we also report adjusted R² (*74*), which penalizes model complexity as penalized R² = 1 − [(1 − R²)(n − 1)/(n − p − 1)], where n is the number of observations used in the prewhitened regression and p is the number of predictors (excluding the intercept). In our primary specification with a single predictor and relatively small monthly blocks, R² provides an intuitive measure of variance explained, while penalized R² was preferred for comparing specifications and guarding against overfitting, particularly when exploring lagged terms that increase the ARIMA p component. We present both metrics but rely on penalized R² for model selection across transfer structures; close agreement between R² and penalized R² indicates a parsimonious model with genuine explanatory power, whereas divergence suggests that apparent in-sample fit may be inflated by model complexity relative to sample size.

### Modeling Assumptions

Time-series methods, including TFM, assume stationarity (that the statistical properties of a series (its mean, variance, and autocorrelation) remain stable over time and violating this assumption risks detecting spurious associations driven by shared trends rather than genuine short-run coupling. Both the purity and mortality series are clearly non-stationary over the study period (purity rises then falls, mortality declines) but the differencing component (d=1) of our ARIMA(1,1,0) prewhitening filter explicitly addresses this by transforming both series into their month-to-month changes before estimating the transfer function, effectively removing the non-stationary trend regardless of whether it is deterministic or stochastic. The additional step of dividing the study period into two analytic blocks provides further protection, as any residual non-stationarity from the structural shift between the rising and declining phases is isolated rather than allowed to span the full series.

### Negative Controls

Any set of indicators that share a structurally similar time trajectory (rise through 2021–2022, decline thereafter) should produce a large, statistically significant association regardless of causality. We conducted a negative control cross-correlation analysis using two common macroeconomic indicators (Supplement S2) with no plausible pharmacological pathway to overdose mortality: the Consumer Price Index (CPI, the US Bureau of Labor Statistics (*79*) monthly measure of consumer inflation), and the University of Michigan Consumer Sentiment Index (*80*) (MCSI, a survey-based measure of household economic confidence). For each negative control, we computed cross-correlation functions (CCFs) on unadjusted time-series, and then applied the same TFM process, using prewhitened residuals obtained by fitting auto-ARIMA models.

### Ethics Statement

This study used public, geographically aggregated, anonymized data and were not eligible for human subjects review, also because we used only data from decedents. The study was pre-registered at the Open Science Foundation: https://osf.io/h6fdw/.

### Artificial Intelligence (AI)

Generative artificial intelligence was used to create initial drafts of Python code, deployed within Deepnote.com notebooks. Instead of being one proprietary standalone foundation model, Deepnote AI functions as a virtual collaborator that relies on an ensemble third-party generative AI models; specifically for code generation, Codeium Windsurf was deployed. Generative AI was also used to generate initial drafts of R code for ARIMA models, using Claude Opus 4.7 (Anthropic). All code was reviewed and modified by humans, and final models were run in traditional (non-AI) integrated development environments.

## Data Availability

All data and code used in the analysis are publicly available at 10.5061/dryad.76hdr7t9q

https://doi.org/10.5061/dryad.zkh1893rq

## ACKNOWLEDGEMENTS

We acknowledge the intellectual contribution of Vangelov et al. to open this discussion, and for their innovative methods which we replicated. We thank the peer reviewers who provided material feedback to improve the manuscript. We are grateful to our colleagues at our institutions who provide support services for pre-processing data, as well as accounting and administrative support.

## Author contributions

Conceptualization: ND, AS, AK, RP, PG, LAP

Methodology: ND, LAP, AS, KGC

Data analysis: ND, KGC

Visualization: ND

Supervision: PG

Writing—original draft: ND, AS

Writing—review & editing: ND, AS, AK, RP, LAP, PG, ST, KGC

## Data and materials availability

All data are available in the main text or the supplementary materials. Code, datasets, codebook, and output in the form of Jupyter .ipynb notebooks are available from the Dryad repository: https://doi.org/10.5061/dryad.zkh1893rq

