## Supplemental materials for "Fentanyl Purity and Overdose Decline: A Reexamination of Geographic Trends"

Science Advances
AAAS

Supplementary Materials for

### Fentanyl Purity and Overdose Decline: A Reexamination of Geographic Trends

Nabarun Dasgupta* *et al.*

**This PDF file includes:**

S1: Fentanyl Purity Data Extraction
S2: Negative Macroeconomic Controls
S3: Sampling Frame of 2023 DEA Fentanyl Powder Exhibits
Figs. S1 to S3
Tables S1 to S4

#### S1: Fentanyl Purity Data Extraction

The fentanyl powder purity time series was obtained by digitizing the trend line from the published figure on page 23 of the 2025 DEA National Drug Threat Assessment, replicating the approach described by Vangelov et al. (7). Because the published figure was available only as a rendered image within an electronic document, direct numerical extraction was not possible, and a coordinate-based digitization procedure was employed.

##### *Digitization Method*

Monthly data points along the fentanyl powder purity trend line (red line, top panel of the DEA figure) were identified by visual inspection and plotted as anchor points on a vector path in Adobe Illustrator. Each anchor point was placed at the center of the plotted data marker or at the mid-line position for months where markers were not visually distinct. The path coordinates were then extracted programmatically using a custom JavaScript routine that read the anchor point positions from the Illustrator document object model and scaled them to the axis system of the original figure. The x-axis was mapped to monthly intervals (January 2019 through September 2024), and the y-axis was calibrated to the purity percentage scale using the labeled gridlines in the published figure.

Pixel-level anti-aliasing in the electronic PDF introduced minor positional ambiguity at each data point. To minimize systematic bias from aliasing artifacts, a conservative placement strategy was used: anchor points were positioned at the visually estimated centroid of the anti-aliased region rather than at any single edge, and borderline cases were resolved by erring toward the nearest labeled gridline. This approach may introduce small positive bias in extracted values relative to the true underlying data, as noted in the replication fidelity analysis below.

##### *Replication Fidelity*

The extracted time series was compared with the data published in the GitHub repository accompanying Vangelov et al. (*3*) using three complementary agreement statistics. Lin's concordance correlation coefficient (*5*), which simultaneously measures precision and accuracy between two sets of measurements, was 0.999, indicating near-perfect reproducibility. Bland–Altman analysis (*6*) yielded a mean difference of +0.164 percentage points (our extraction marginally higher than Vangelov et al.), with 95% limits of agreement spanning −0.182 to +0.509 percentage points. Pearson correlation between the two extractions was $r > 0.999$. These metrics collectively demonstrate negligible inter-extraction disagreement across all monthly observations (Fig. S1).

##### *Coordinate Extraction Code*

The custom JavaScript used for Illustrator coordinate extraction and axis read anchor point positions from a selected path object, applies linear interpolation to map pixel coordinates to the figure's axis units, and outputs a CSV of monthly purity values.

We note that the pill purity data from an adjacent panel in the same DEA figure were not re-analyzed. No observable correlation existed between pill and powder purity trends (Fig. S2), the magnitude of pill concentration variation was substantially lower, and irregular axis labeling (the vertical axis did not include zero) raised concerns about the validity of manual extraction for that series.

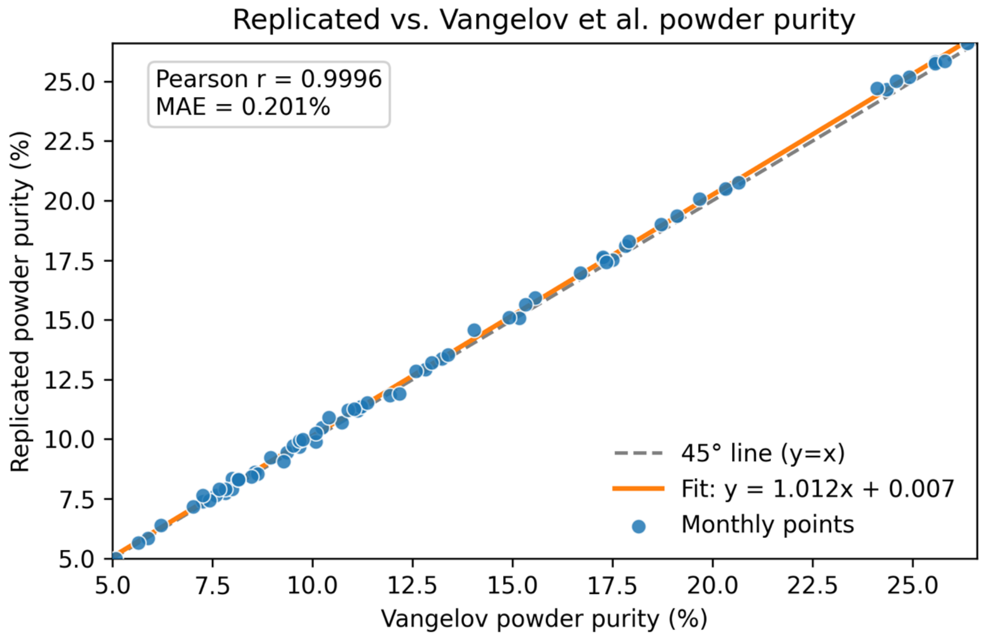


**Fig. S1. Replication fidelity of fentanyl powder purity data extraction.** Comparison of monthly fentanyl powder purity values extracted independently in the present study versus those published in the Vangelov et al. GitHub repository (*3*). Lin's concordance correlation coefficient = 0.999; Bland–Altman mean difference = +0.164 percentage points (limits of agreement: −0.182 to +0.509). The close agreement confirms that the digitization procedure reproduced the original data with negligible measurement error.

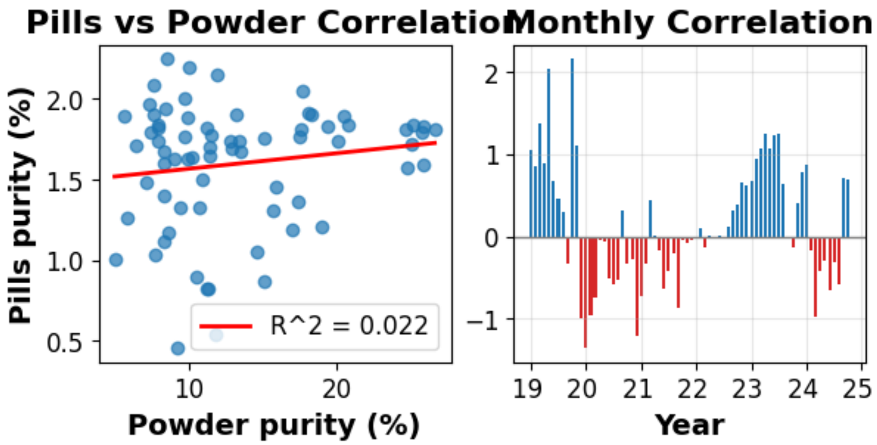


**Fig. S2. Correlation between pill and powder fentanyl potency reported by DEA.** Scatterplot of monthly DEA-reported fentanyl pill potency (milligrams per tablet) versus powder purity (percent) across the study period. No meaningful correlation was observed between the two series, supporting the decision to restrict the primary analysis to powder purity data.

In our primary analysis, we did not include pill purity data from an adjacent figure in the DEA report because there was no observable correlation with powder purity, pills had a much lower magnitude of concentration, and irregular labeling of the figure's vertical axis (e.g., fails to include zero) raised questions about the validity of manual data extraction.

#### S2: Negative Macroeconomic Controls

##### *Rationale*

A central methodological concern in interpreting the analysis by Vangelov et al. is that the temporal correlation between nationally aggregated DEA fentanyl purity and overdose mortality may reflect shared post-COVID macrotrends rather than a pharmacologically meaningful supply-shock signal. Both fentanyl purity and overdose mortality followed a characteristic trajectory over 2019–2024: relative stability in the pre-pandemic period, disruption and surge during 2020–2021, peak plateau in 2022–2023, and concurrent decline thereafter. This trajectory is not unique to the drug supply or drug-related mortality—it characterizes a broad range of social and economic indicators affected by pandemic disruption and recovery.

When two non-stationary time series share a common trend, standard correlation statistics will detect a strong association even in the complete absence of any causal or mechanistic relationship. This phenomenon, known as spurious correlation, is well described in the time-series literature and motivates autocorrelation-adjustment procedures such as Box-Jenkins prewhitening prior to cross-correlation analysis.

To illustrate the magnitude and direction of this confounding in the present context, we conducted a supplementary cross-correlation analysis using two economic indicators—the Consumer Price Index (CPI) and the University of Michigan Consumer Sentiment Index—that have no plausible direct pharmacological pathway to overdose mortality. If the unadjusted correlation between DEA purity and overdose mortality is meaningful, it should be substantially stronger than correlations with causally unrelated series. If similarly large correlations arise with multiple causally unrelated indicators, and if these correlations attenuate or reverse sign following autocorrelation adjustment, this provides strong evidence that the unadjusted purity–mortality association is driven primarily by shared post-COVID trend structure.

##### *Methods*

###### *Data Sources and Variables*

Monthly data spanning January 2019 through September 2024 were assembled from the sources listed in Table S1.

**Table S1.** Data sources, date ranges, and units for all variables included in the supplementary cross-correlation analysis.

| Variable | Source | Date Range | Units |
|---|---|---|---|
| OD Mortality Rate | NCHS/CDC NVSS via OD Pulse | Jan 2019–Sep 2024 | Deaths per 100,000 population/month |
| Consumer Price Index | U.S. Bureau of Labor Statistics | Jan 2019–Sep 2024 | 12-month % growth rate |
| Michigan Consumer Sentiment | Univ. of Michigan Survey | Jan 2019–Sep 2024 | Index (1966 = 100) |

*Analytical Approach*

All analyses were conducted in R (version 4.3.3) using the *stats*, *forecast*, *ggplot2*, and *patchwork* packages. The analysis proceeded in three stages.

*Stage 1: Visual Co-movement (Unadjusted Levels)*

Pearson correlation coefficients were computed between the OD mortality rate (levels) and each economic indicator (levels) to quantify the unadjusted association. Series were Z-score normalized (mean = 0, SD = 1) and plotted together on a shared time axis. Pearson $r$ was computed on the original unstandardized series.

*Stage 2: Unadjusted Cross-Correlation Function*

Cross-correlation functions (CCFs) were computed between OD mortality rate and each economic indicator across lags of −12 to +12 months, without autocorrelation adjustment. In the CCF, a positive lag $k$ indicates that the input series at time $t$ is associated with the output series at time $t+k$. The 95% confidence interval was computed as $\pm 1.96/\sqrt{n}$, where $n = 69$. A CCF in which nearly all bars exceed the 95% CI across all lags is the expected signature of two autocorrelated, trending series and does not indicate genuine dynamic coupling.

*Stage 3: Box-Jenkins Prewhitening*

To remove autocorrelation structure from both series prior to CCF estimation, a standard Box-Jenkins prewhitening procedure was applied (*7, 8*). An ARIMA model was fitted to the OD mortality rate series (the input series) using automatic model selection via *auto.arima()* with Bayesian Information Criterion (BIC) and full model search (stepwise = FALSE). The selected model was ARIMA(0,2,1)(1,0,0)[12], indicating that the OD rate series required second-order differencing to achieve stationarity, with a moving-average term at lag 1 and a weak seasonal autoregressive component.

The residuals of this ARIMA fit constituted the prewhitened input series ($\alpha_t$). The same AR/I/MA filter structure (order $p = 0$, $d = 2$, $q = 1$, seasonal $P = 1$, $D = 0$, $Q = 0$) was applied to the economic indicator series using the *Arima()* function with identical order constraints, yielding prewhitened output residuals ($\beta_t$). CCF was then computed between $\alpha_t$ and $\beta_t$ across lags −12 to +12 months.

**Table S2.** Auto-ARIMA models selected by BIC for each series. The OD mortality rate filter (ARIMA(0,2,1)(1,0,0)[12]) was applied uniformly to both economic indicator series during prewhitening.

| Series | Auto-ARIMA Model (BIC) | AICc | Interpretation |
|---|---|---|---|
| OD Mortality Rate | ARIMA(0,2,1)(1,0,0)[12] | −95.5 | Second-order integrated; strong non-linear trend with weak seasonal AR |
| CPI (12-mo %) | ARIMA(1,1,0)(1,0,0)[12] | 60.5 | First-order integrated with AR(1) inertia and seasonal AR |
| Sentiment Index | ARIMA(0,1,0) | 414.9 | Random walk after first differencing; no exploitable autocorrelation |

***Results***

*Unadjusted Associations*

In unadjusted levels analysis, the OD mortality rate exhibited large correlations with both economic indicators (Table S3). The OD rate was positively correlated with CPI (Pearson $r$ = +0.63, contemporaneous) and negatively correlated with the Consumer Sentiment Index ($r$ = −0.73, lag −1 month; sentiment leading OD rate). The CCFs for both pairs showed the characteristic pattern of spurious correlation: bars significant across virtually all lags from −12 to +12 months, with no distinguishable peak lag structure (Fig. S3, Rows B1 and B2). This pattern is expected when two autocorrelated, trending series are compared without adjustment and does not constitute evidence of meaningful dynamic coupling.

*Prewhitened Associations*

After Box-Jenkins prewhitening, both associations changed substantially in magnitude and direction (Table S3; Fig. S3, Rows C1 and C2). The OD rate–CPI correlation reversed sign (prewhitened peak $r$ = −0.34 at lag +1 month; unadjusted $r$ = +0.63), indicating that the positive unadjusted correlation was entirely attributable to shared trend structure. The direction reversal—from positive to negative—cannot be explained by residual confounding or lag misspecification; it demonstrates that when shared trends are removed, the innovations of CPI and OD mortality move in opposite directions.

The OD rate–Sentiment correlation also reversed sign (prewhitened peak $r$ = +0.32 at lag +3 months; unadjusted $r$ = −0.73 at lag −1 month). The original negative association reflected the concurrent 2020–2022 deterioration and 2023–2024 recovery in both indicators. After prewhitening, the residual positive association at lag +3 months likely reflects a COVID-period artifact in which the overdose surge slightly lagged the initial lockdown shock that had already begun driving a sentiment recovery.

**Table S3.** Peak cross-correlation coefficients before and after Box-Jenkins prewhitening.

| Series pair | Unadjusted r | Unadj. lag | Prewhitened r | Prewhit. lag |
|---|---|---|---|---|
| OD Rate ~ CPI | +0.63* | 0 | **−0.34*†** | +1 |
| OD Rate ~ Sentiment | −0.73* | −1 | **+0.32*†** | +3 |

** $p < 0.05$ ($|r| > 1.96/\sqrt{n} = 0.236$, $n = 69$). † Sign reversal relative to unadjusted estimate.*

##### *Interpretation*

The central finding of this supplementary analysis is that two economic indicators with no pharmacological mechanism linking them to overdose mortality produce unadjusted correlations with the OD mortality rate ($|r| = 0.63–0.73$) that are comparable to or larger than the correlation reported by Vangelov et al. between DEA fentanyl purity and overdose mortality. Both correlations are attributable to the shared post-COVID trend: a broad rise in multiple social and economic stressors through 2021–2022, followed by concurrent amelioration from 2023 onward.

This does not imply that the purity–mortality relationship is necessarily spurious in every context. Rather, it demonstrates that in the presence of strong, shared non-stationarity of the kind observed across multiple social metrics in the 2019–2024 period, an unadjusted correlation between any two trending series will appear large and statistically significant regardless of whether any genuine relationship exists. The appropriate analytic response—as implemented in the transfer function models reported in the main text—is to remove the autocorrelation structure from both the input (purity) and output (mortality) series before estimating dynamic associations, so that inference is based on month-to-month innovations rather than shared long-run trends.

Fentanyl purity also followed the same broad post-COVID arc (rising through 2022, declining thereafter), making it structurally identical, from a time-series perspective, to CPI and Consumer Sentiment. An analysis that does not account for this shared trend cannot distinguish a causal supply-shock signal from the kind of spurious co-movement illustrated here.

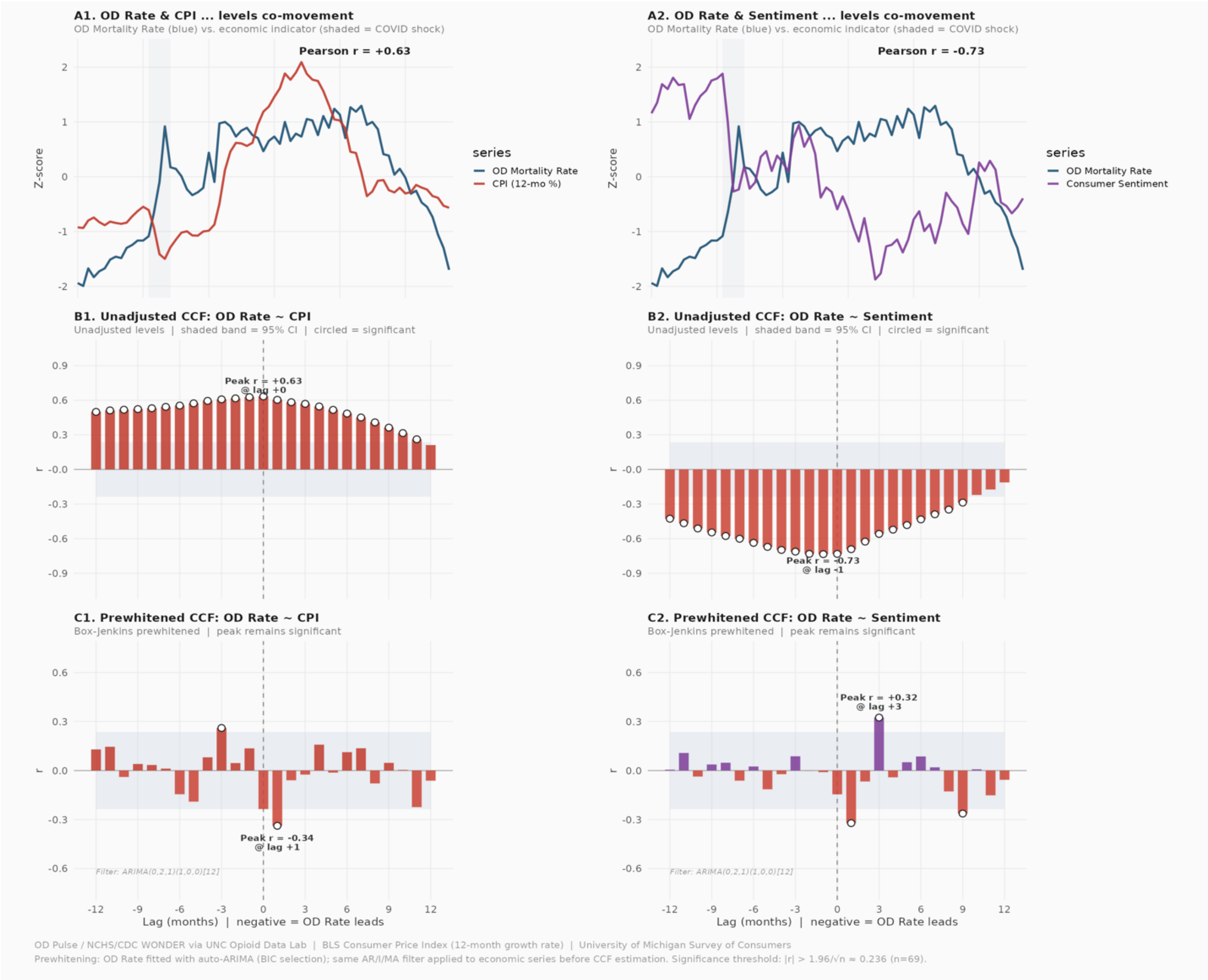


**Fig. S3. Cross-correlation analysis of overdose mortality versus macroeconomic indicators.**
Row A: Z-score normalized time series of OD mortality rate (red) overlaid with each economic indicator (blue) showing visual co-movement. Row B: Unadjusted cross-correlation functions (CCFs) across lags −12 to +12 months; virtually all bars exceed the 95% confidence interval (dashed lines), the expected signature of spurious correlation between autocorrelated trending series. Row C: Prewhitened CCFs after Box-Jenkins filtering; both associations reverse sign and attenuate markedly, demonstrating that the unadjusted correlations were artifacts of shared post-COVID trend structure rather than genuine dynamic coupling.

#### S3: Sampling Frame of 2023 DEA Fentanyl Powder Exhibits

##### *DEA's Two Purity Data Sources: Regional Field Laboratories vs. the Fentanyl Profiling Program*

DEA publicly reports fentanyl purity data from two distinct laboratory systems with different sampling frames and analytic purposes (35). Understanding their differences is essential for interpreting any purity time series derived from DEA data.

The first source is the network of 10 DEA regional field laboratories distributed across the United States, where routine law enforcement seizures—ostensibly at or near the street retail level—are identified and quantitated within the context of criminal prosecution. In CY [calendar year] 2023, $n$ = 7,857 fentanyl powder samples were tested across these regional. The geographic distribution of exhibits across states, presented in Table S4 below, is drawn from this source. For CY 2023, the average powder purity reported by these regional field laboratories was 19.7%.

The second source is the DEA's Fentanyl Profiling Program (FPP), housed within the Special Testing and Research Laboratory (SFL1) in the Office of Forensic Sciences. FPP conducts detailed forensic chemical analysis on a smaller, intentionally selected subset of seizures, including synthetic route classification, adulterant profiling, and purity quantitation for scientific and intelligence purposes.

In recent years, FPP shifted its sampling focus toward higher-volume ("wholesale") seizures. DEA (35) explicitly stated:

> *"The significant increase in purity in CY [calendar year] 2023 is likely due to a change in the FPP sampling plan, which changed to focus more on wholesale level seizures. These larger seizures generally have a higher purity as they are not yet significantly adulterated domestically."*

The report further specified the timing:

> *"In January 2023, FPP focused efforts on seized samples weighing greater than 800 g. In July 2023, new sampling guidance was released to the DEA laboratory system advising the submission of samples containing fentanyl, fentanyl-related compounds, and precursor chemicals when the net weight is greater than or equal to 800 g."*

For CY 2023, the average powder purity reported by FPP was 33.5%—nearly 70% higher than the 19.7% average from the regional field laboratories. This discrepancy is a direct and expected consequence of the 800-gram minimum weight threshold, which systematically selects for wholesale-level seizures that have not yet been adulterated for retail distribution.

The composition of the purity time series published in the 2025 National Drug Threat Assessment (the series digitized and analyzed by Vangelov et al.) was not specified in that report. It is unclear whether this series includes FPP wholesale samples, regional field lab retail samples, or both, or whether the two sources are statistically weighted. The July 2023 sampling guidance was also provided to the regional DEA field laboratories, further complicating interpretation of any aggregate purity estimate from this period. Discrepancies between the Vangelov et al. purity values and other published DEA reports (discussed in the main text) suggest that higher-purity wholesale FPP samples were comingled with routinely collected retail-level field laboratory samples, potentially inflating the apparent purity peak in 2023.

***Putative sampling frame***

We are able to provide context (Supplement S3) for the sampling distribution of fentanyl powder samples analyzed by DEA regional labs. Out of 7,737 total samples analyzed in 2023, the South accounted for 3,370 (43.56%), followed by the Northeast with 1,987 (25.68%), the Midwest with 1,263 (16.32%), and the West with 1,117 (14.44%).

Three states (Florida n=751, California n=657, and New York n=498) made up 24.6% of exhibits nationwide. Within the South, Florida accounted for 22.3% of samples, followed by West Virginia (n=459, 13.6%). Within the Midwest, Missouri (n=413, 32.7%) and Illinois (n=229, 18.1%) are the top states by exhibit volume. And in the West it was California (n=657, 58.8%) and Washington (13.1%). In the Northeast, New York (n=498, 25.1%) and Massachusetts (n=339, 17.1%) were the top two. These distributions aid in interpretation of regionally stratified TFM models.

These distributions should be considered when interpreting the “national” fentanyl purity series. Net seizure weight does not correspond proportionally to exhibit counts; for example, Tennessee contributed 219 exhibits representing 223.2 kg, while West Virginia contributed 459 exhibits representing only 12.3 kg, reflecting differences in seizure size distributions across jurisdictions.

***Geographic Distribution of Regional Field Laboratory Exhibits***

Table S4 presents the geographic distribution of fentanyl powder exhibits analyzed by the DEA regional field laboratories (not the FPP) in CY 2023. These data are drawn from the AIDD powders table on page 4 of the Annual Fentanyl Report CY 2023.

**Table S4.** Fentanyl powder exhibits analyzed by DEA regional field laboratories (not the Fentanyl Profiling Program), by state and U.S. Census Region, calendar year 2023. Source: DEA *Annual Fentanyl Report CY 2023* (PRB# 2025-017), AIDD data, page 4 (powders column)

| State | Census Region | No. Exhibits | % Natl. | % Region | Net Wt. (kg) | % Natl. Wt. | % Reg. Wt. |
|---|---|---|---|---|---|---|---|
| CT | Northeast | 301 | 3.9 | 15.1 | 28.4 | 1.3 | 5.8 |
| MA | | 339 | 4.4 | 17.1 | 84.3 | 3.8 | 17.2 |
| ME | | 71 | 0.9 | 3.6 | 8.4 | 0.4 | 1.7 |
| NH | | 119 | 1.5 | 6.0 | 13.6 | 0.6 | 2.8 |
| NJ | | 255 | 3.3 | 12.8 | 60.0 | 2.7 | 12.2 |
| NY | | 498 | 6.4 | 25.1 | 227.7 | 10.2 | 46.4 |
| PA | | 293 | 3.8 | 14.7 | 48.6 | 2.2 | 9.9 |
| RI | | 49 | 0.6 | 2.5 | 16.9 | 0.8 | 3.4 |
| VT | | 62 | 0.8 | 3.1 | 2.6 | 0.1 | 0.5 |
| | **NE Subtotal** | **1987** | **25.7** | **100.0** | **490.5** | **22.0** | **100.0** |
| IA | Midwest | 15 | 0.2 | 1.2 | 0.5 | 0.0 | 0.2 |
| IL | | 229 | 3.0 | 18.1 | 107.1 | 4.8 | 35.0 |
| IN | | 139 | 1.8 | 11.0 | 30.9 | 1.4 | 10.1 |
| KS | | 8 | 0.1 | 0.6 | 11.9 | 0.5 | 3.9 |
| MI | | 167 | 2.2 | 13.2 | 74.3 | 3.3 | 24.3 |
| MN | | 2 | 0.0 | 0.2 | 0.04 | 0.0 | 0.0 |
| MO | | 413 | 5.3 | 32.7 | 60.3 | 2.7 | 19.7 |
| ND | | 4 | 0.1 | 0.3 | 0.1 | 0.0 | 0.0 |
| NE | | 15 | 0.2 | 1.2 | 0.1 | 0.0 | 0.0 |
| OH | | 158 | 2.0 | 12.5 | 14.0 | 0.6 | 4.6 |
| SD | | 1 | 0.0 | 0.1 | 0.01 | 0.0 | 0.0 |
| WI | | 112 | 1.4 | 8.9 | 7.0 | 0.3 | 2.3 |
| | **MW Subtotal** | **1263** | **16.3** | **100.0** | **306.3** | **13.7** | **100.0** |
| AL | South | 96 | 1.2 | 2.8 | 47.1 | 2.1 | 5.6 |
| AR | | 73 | 0.9 | 2.2 | 13.5 | 0.6 | 1.6 |
| DC | | 179 | 2.3 | 5.3 | 4.6 | 0.2 | 0.5 |

| | | | | | | | |
|---|---|---|---|---|---|---|---|
| DE | | 78 | 1.0 | 2.3 | 12.3 | 0.6 | 1.5 |
| FL | | 751 | 9.7 | 22.3 | 167.2 | 7.5 | 19.9 |
| GA | | 154 | 2.0 | 4.6 | 25.5 | 1.1 | 3.0 |
| KY | | 209 | 2.7 | 6.2 | 31.4 | 1.4 | 3.7 |
| LA | | 54 | 0.7 | 1.6 | 11.7 | 0.5 | 1.4 |
| MD | | 129 | 1.7 | 3.8 | 14.6 | 0.7 | 1.7 |
| MS | | 33 | 0.4 | 1.0 | 3.6 | 0.2 | 0.4 |
| NC | | 296 | 3.8 | 8.8 | 59.9 | 2.7 | 7.1 |
| OK | | 115 | 1.5 | 3.4 | 27.0 | 1.2 | 3.2 |
| PR | | 95 | 1.2 | 2.8 | 6.9 | 0.3 | 0.8 |
| SC | | 88 | 1.1 | 2.6 | 48.0 | 2.2 | 5.7 |
| TN | | 219 | 2.8 | 6.5 | 223.2 | 10.0 | 26.5 |
| TX | | 215 | 2.8 | 6.4 | 109.2 | 4.9 | 13.0 |
| VA | | 127 | 1.6 | 3.8 | 23.1 | 1.0 | 2.7 |
| WV | | 459 | 5.9 | 13.6 | 12.3 | 0.6 | 1.5 |
| | **South Subtotal** | **3370** | **43.6** | **100.0** | **841.1** | **37.7** | **100.0** |
| AK | West | 18 | 0.2 | 1.6 | 3.7 | 0.2 | 0.6 |
| AZ | | 67 | 0.9 | 6.0 | 66.0 | 3.0 | 11.2 |
| CA | | 657 | 8.5 | 58.8 | 416.1 | 18.7 | 70.3 |
| CO | | 27 | 0.3 | 2.4 | 8.6 | 0.4 | 1.5 |
| HI | | 23 | 0.3 | 2.1 | 3.4 | 0.2 | 0.6 |
| ID | | 3 | 0.0 | 0.3 | 0.03 | 0.0 | 0.0 |
| MT | | 38 | 0.5 | 3.4 | 1.4 | 0.1 | 0.2 |
| NM | | 9 | 0.1 | 0.8 | 5.1 | 0.2 | 0.9 |
| NV | | 19 | 0.2 | 1.7 | 1.7 | 0.1 | 0.3 |
| OR | | 109 | 1.4 | 9.8 | 50.1 | 2.2 | 8.5 |
| UT | | 1 | 0.0 | 0.1 | 0.01 | 0.0 | 0.0 |
| WA | | 146 | 1.9 | 13.1 | 35.6 | 1.6 | 6.0 |
| | **West Subtotal** | **1117** | **14.4** | **100.0** | **591.7** | **26.5** | **100.0** |

| | | | | | | | |
|---|---|---|---|---|---|---|---|
| | **National Total** | **7737** | **100.0** | — | **2229.6** | **100.0** | — |

**Notes:** Exhibits reflect powder forms only (crystalline, powder, and rock-like); tablet exhibits ($n$ = 5,958) are excluded. Percentages may not sum to exactly 100.0% due to rounding. Census Region assignments follow U.S. Census Bureau definitions. DC and PR are classified within the South following DEA field office jurisdictional convention. The DEA regional laboratory system comprises ten laboratories distributed across the United States; exhibit counts per state reflect the volume of drug evidence submitted to and analyzed by the laboratory serving each jurisdiction, not a statistically representative sample of the illicit drug supply. The regional field laboratory average powder purity for CY 2023 was 19.7%, compared to 33.5% for the intentionally selected FPP wholesale samples.
